# The importance of adolescent girls and “*epidemic gearing*” on HIV prevalence across West Africa

**DOI:** 10.1101/19008839

**Authors:** Holly J Prudden, Zindoga Mukandavire, Marelize Gorgens, David Wilson, Jasmina Panovska-Griffiths, Charlotte Watts

## Abstract

**Background:** In West Africa HIV prevalence varies between 0.1-6% in female and between 0.1-4% in the male general population. Male circumcision is almost universal, and it is unclear what drives this variation. We use mathematical modelling to identify the determinants of this variation across fourteen West African countries.

**Methods:** We developed a novel dynamic model of HIV transmission between population cohorts of female sex workers (FSWs), their clients, females with 2+ partners in the past year and other sexually active women and men in the general population. Parameter ranges were determined from the literature and sampled using Latin Hypercube sampling to identify parameter sets that fit West African HIV prevalence data. Partial-rank correlation coefficients between different model parameters and the HIV prevalence in general male and female population across 14 countries were calculated to determine to most significantly correlated model parameters to HIV prevalence.

**Results:** The key determinant of HIV in females when prevalence is between 0-3% is the size of the brothel and non-brothel FSW groups. When female HIV prevalence >3%, the percentage of sexually active adolescent females with 2+ partners has greater influence on HIV prevalence. The size of the FSW groups has the most significant impact on HIV prevalence for males.

**Conclusions:** Our findings confirm the role of FSWs in West Africa as an important determinant of HIV risk, but also identify, in countries with higher HIV prevalence, the emerging role of a group of adolescent girls with 2+ partners is an important determinant of risk. In fact, our findings suggest that this group may enable the epidemic to be effectively “*geared up”* when partnerships are formed with higher-risk males, indicating additional prevention needs amongst this group.

**Funding:** This study was funded by UNAIDS.

## Introduction

HIV in West Africa accounts for around 20% of all infections in sub-Saharan Africa with HIV prevalence here generally lower than in other African regions [1] most likely as a result of the almost universal practice of male circumcision across West Africa [2]. Around 30% of all AIDS-related deaths worldwide occur in West Africa with reported 280,000 AIDS-related deaths in 2017 [3]but are significant variations in HIV prevalence between countries (Figure 1). The National Demographic and Health Surveys (DHS) suggest that Nigeria, Cote d’Ivoire and Cameroon have the highest HIV prevalence levels (4-6% among females and 2-4% in males) in comparison to other countries (0.5-3% amongst both females and males [4,5]

**Figure 1.**
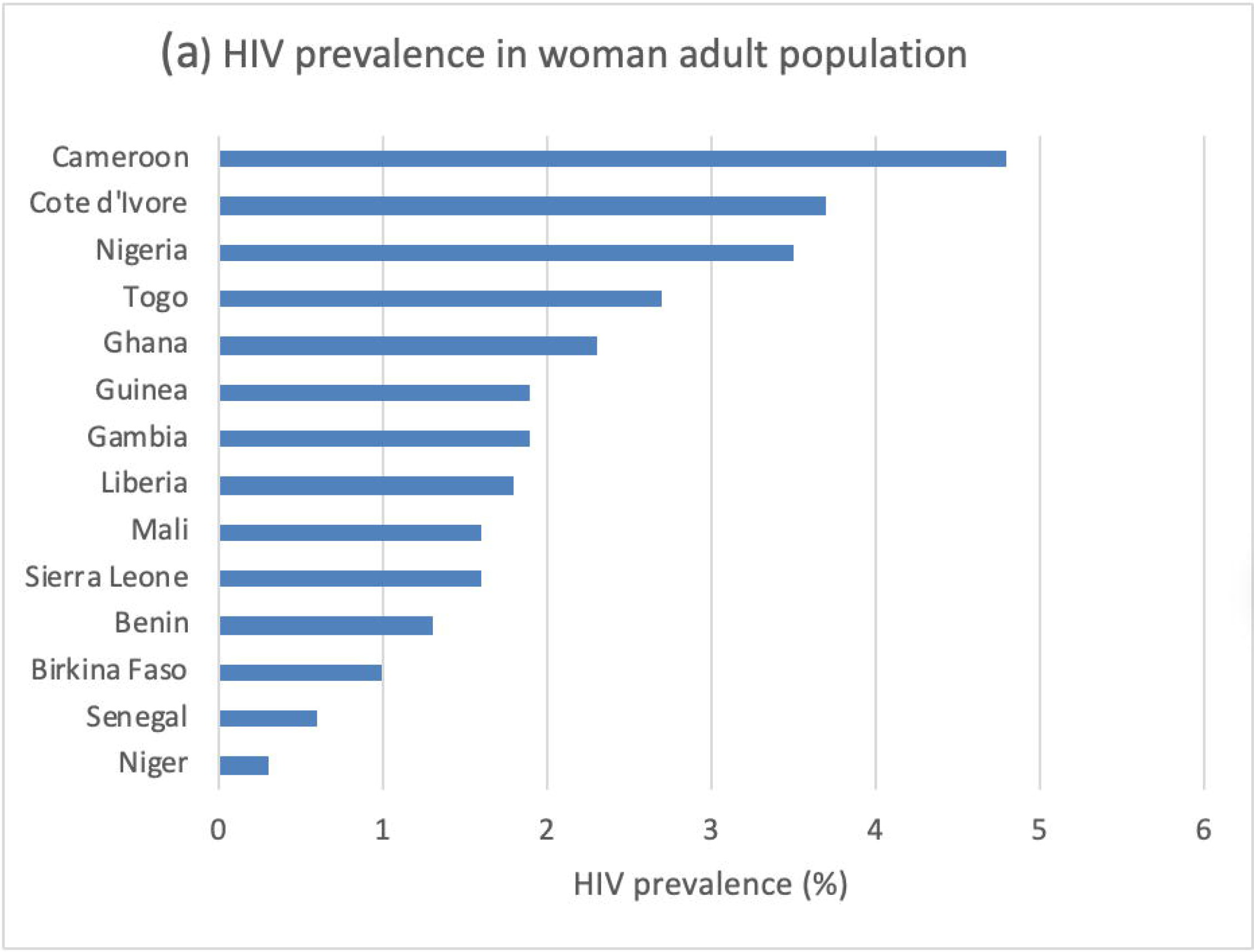
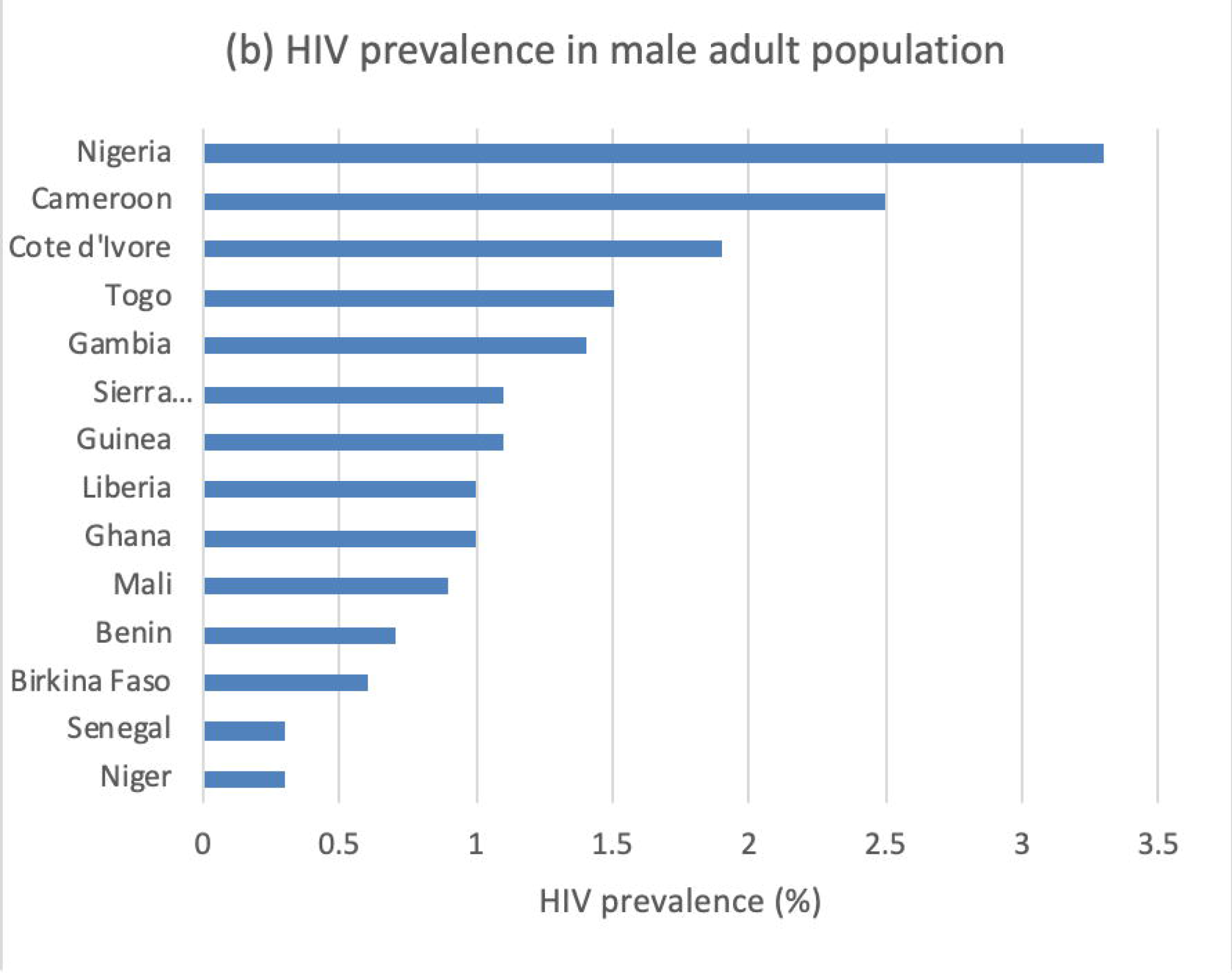
(a) and (b) HIV prevalence in the general adult female (a) and male (b) populations across 14 West African countries. HIV prevalence data is taken from UNAIDS report https://www.avert.org/hiv-and-aids-west-and-central-africa-overview from 2017

Recent UNAIDS reports suggest that women are disproportionately affected by HIV in West and Central Africa and account for around 57% of adults living with HIV in 2017. The average HIV prevalence stands at 2.3% among adult women (15-49 years old), compared to 1.6% among adult men (15-49 years old) [4]. In addition, reports suggest that adolescent girls and young women (aged 15-24) are disproportionately more likely to acquire HIV than their male counterparts [4].

Past epidemiological studies from this region have suggested that commercial sex is a major driver of the HIV epidemic [5] and that interventions focused on female sex workers (FSWs) and their clients are highly effective at reducing the prevalence of sexually transmitted infections (STIs) [5]. In addition, a mathematical modelling study has suggested commercial sex work as a major driver of both the short and longer term epidemic trajectories in the region [6]. However, our ecological analysis of variations in population prevalence levels across West Africa highlight that, along with female commercial sex work, variations in HIV prevalence amongst general population males and females between countries may be associated with national variations in the percentage of younger females (15-24 years) that have 2 or more (2+) partners in the past year, as defined by Demographic Health surveys [7]. The role of this adolescent girls group has previously been mentioned as a key population for HIV control [8].

Our work expands this idea and aims to explore the role of this group of younger females (15-24 years) that have 2 or more (2+) partners, in describing the variations in HIV prevalence among different countries in West Africa. The overall aim of this work is to identify the potential determinants of population variation in HIV prevalence among the female and male general population in West Africa, considering 14 countries in the region. Unlike our previous work [7], that focused on ecological analysis, here, we developed and applied a dynamic model for HIV transmission between different cohorts at risk of acquiring HIV in West Africa. Based on existing literature and work to date, we included cohorts of FSW, clients and the general population. The novelty of our work is that, unlike previous studies, within our model we incorporated a cohort of sexually active adolescent females with multiple (2+) partners in the past year and allowed the model to have a proportionate mixing pattern rather than a standard one. This means sexual partnerships between females 2+, clients and males 2+ were distributed proportionately based on their number of partners (as opposed to having a fixed number of sexual partners for each group). Including this group of females with 2+ partners and the proportionate mixing pattern within a dynamic transmission model, allowed us to model the groups’ interactions with other population cohorts and hence explore their role in describing the variations of HIV prevalence in West Africa.

## Methods

The methodology for this work is described in Figure 2(a) with the novel proportionate mixing pattern illustrated in Figure 2(b).

**Figure 2.**
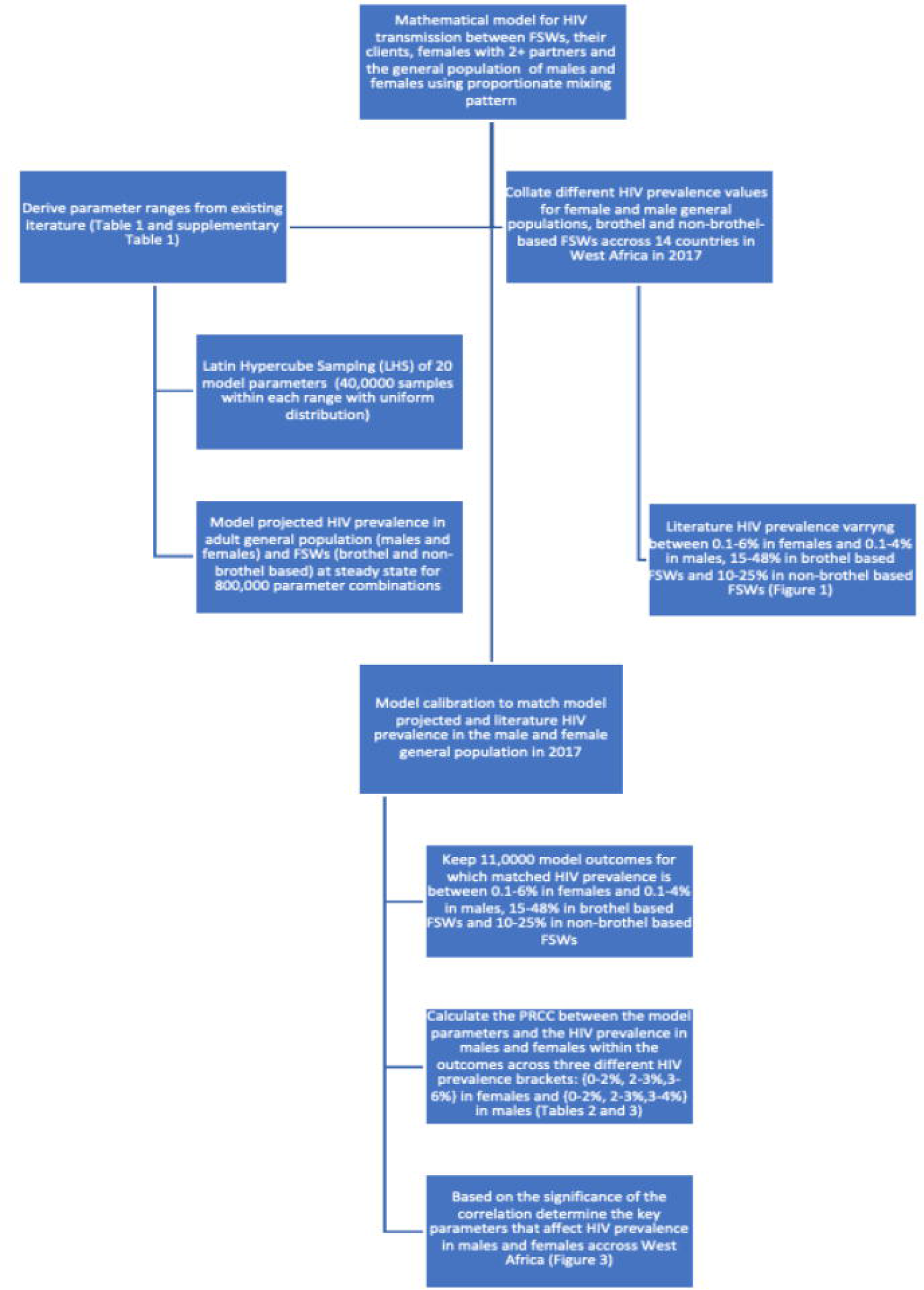
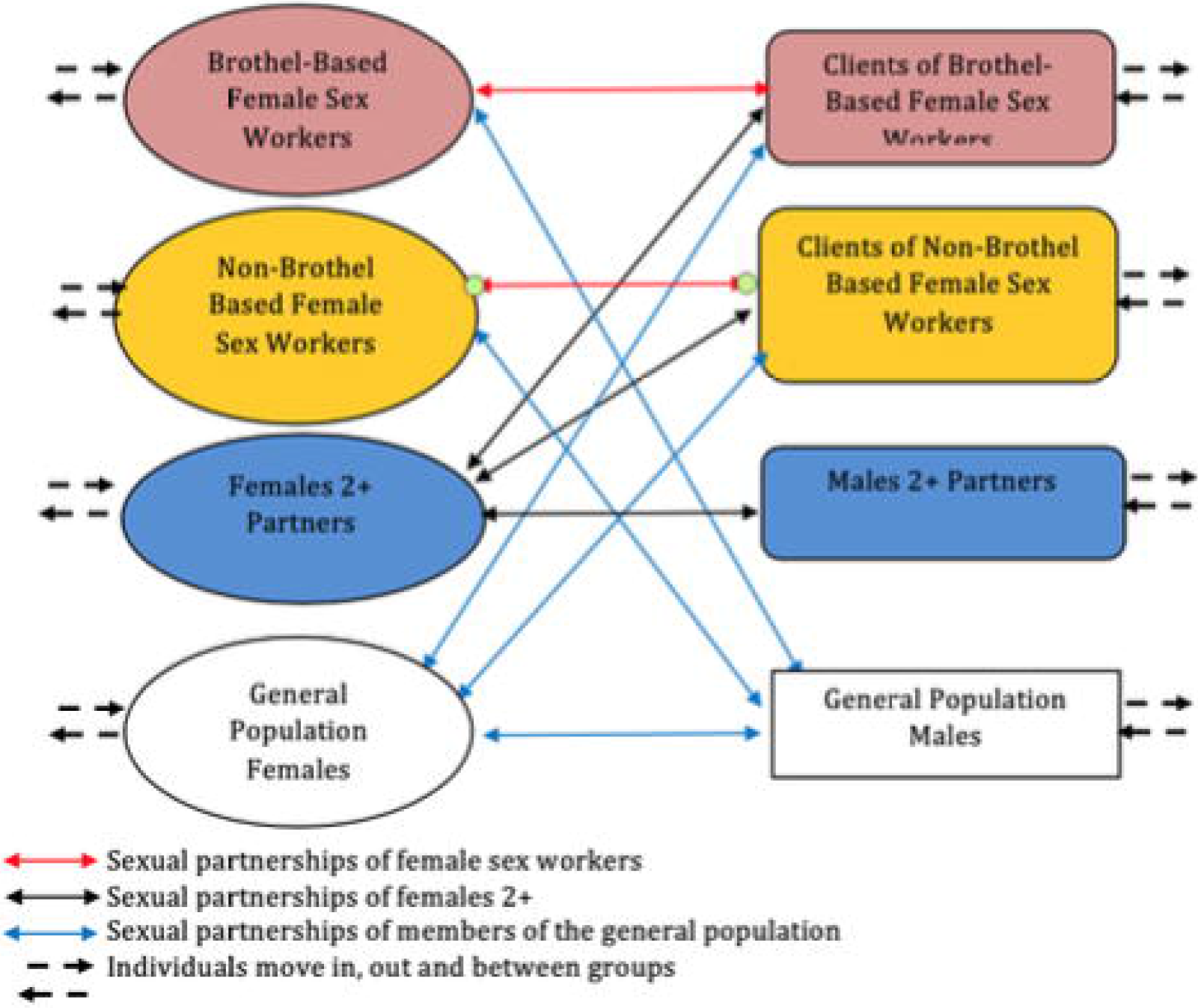
(a): Description of the methodology for this paper. (b) Sexual mixing patterns in the population. Black arrow denote partnership: Sexual partnerships exist between the two FSW groups and their respective client groups and between clients and females with 2+ partners. Younger females 2+ also partner with males 2+. Separate groups of general population males and females form partnerships with one another, with a given proportion also having longer term partnerships with clients (in the case of females) and FSW (in the case of males (to represent the wives of clients and husbands of sex workers, respectively).

### Model design

We developed a deterministic compartmental model describing the transmission of HIV infection among sexually active 15-49 year olds. The mathematical framework and model equations are detailed in Appendix 1. In summary, the model stratifies the sexually active population into two groups of female sex workers (brothel-based and non-brothel based) and their respective client partner groups. In addition, the model includes a separate group of adolescent females with 2+ non-commercial sexual partnerships in the past year and a group of males (15-49) with 2+ non-commercial sex worker partners in the past year, as well as subgroups representing other sexually active men and women in the general population. In order to explore the population sexual mixing pattern that best represents West Africa, we introduced a mixing matrix based on a “fixed proportionate” mixing scenario (depicted in Figure 2(b)). Here, we created an additional parameter, ζ, to represent the proportion of partnerships females 2+ have with men who are also clients (of either brothel-based or non-brothel based FSW). This parameter was also sampled in the course of the uncertainty analysis.

### Model Parameterisation

Biological, epidemiological parameter data and published developmental indicators for West Africa were extracted from the literature. A full parameter table, including additional information on the computation and extraction of data is provided in Appendix 1. Table 1 below is a shortened version of the table showing only the parameters that were used as model inputs for our statistical uncertainty analysis. Based on the data from DHS surveys across the region we assume the entire male population are circumcised [9]. The probability of transmission per act is estimated using a wide range and this incorporates the potential presence of sexually transmitted infections, anal sex acts (in addition to vaginal sex acts) and the effects of anti-retroviral therapy, which is introduced into the model 20 years after the start of the epidemic.

**Table 1:** Biological and behavioural parameter estimates and ranges for countries across the West Africa region with ranges derived from existing literature.

| Description | Symbol | Range of parameter values with reference [X] |
| --- | --- | --- |
| Probability of HIV transmission | $(\beta_{mf}, \beta_{fm})$ | 0.006-0.06 |
| Relative size of Brothel-based FSWs group | $P_{FBB}$ | 0-1.1% |
| Relative size of Non-brothel based FSWs group | $P_{FNB}$ | 0.15-3% |
| Relative size of Females with 2+ partners group | $P_{FTS}$ | 0.1-20% |
| Number of client partners of brothel-based female sex worker. | $C_{FBB}^C$ | 252-1092/yr |
| Number of client partners of non-brothel based FSWs | $C_{FNB}^C$ | 42-336/yr |
| Number of partners of Females 2+ partners (these include client partners of both brothel-based and non-brothel based FSWs as well as males 2+ partners) | $C_{CTS}^{TS}$ | 3-24/yr<sup>1</sup> |
| Number of brothel based FSWs, clients of brothel-based FSW form partnerships with. | $C_{CBB}^C$ | 12-48/yr |
| Number of non-brothel based FSWs, clients of non-brothel based FSWs form partnerships with. | $C_{CNB}^C$ | 12-48/yr |
| Consistency of condom use among Brothel-based female sex workers | $f_{FBB}$ | 41-83% (adjusted for efficacy) |
| Consistency of condom use among Non-brothel based female sex workers | $f_{FNB}$ | 41-68% |
| Consistency of condom use of females with 2+ partners | $f_{FTS}$ | 10-39% |
| Duration of brothel based female sex workers | $1/\alpha_{BB}$ | 0.5-6 yrs |
| Duration of non-brothel based female sex workers | $1/\alpha_{NB}$ | 0.5-6 yrs |
| Duration of Females 2+ | $1/\alpha_{TS}$ | 1-15 yrs |
| Duration of brothel based clients | $1/\varphi_{BB}$ | 5-10yrs |
| Duration of non-brothel based clients | $1/\varphi_{NB}$ | 5-10yrs |
| Duration of Males 2+ | $1/\varphi_{TS}$ | 5-20yrs |
| Percentage of sex acts of females 2+ with client partners of brothel-based and non-brothel based FSWs | $\xi$ | 0-100% |
| (Percentage of sex acts of females 2+ with non-client male partners) | $1 - \xi$ | 0-100% |

### Uncertainty analysis and model calibration

For each of these parameters in Table 1, we generated a range of plausible estimates from the literature to account for the parameter variations across West African countries. We then sampled within each parameter range (splitting it uniformly into 40,000 samples) using a Latin Hypercube sampling [10] to generate 800,000 parameter combinations as inputs to the mathematical model. For each parameter combination, the model was solved numerically in R programming environment until HIV steady state across the different population cohorts was achieved. To calibrate the model, only the projections where HIV prevalence was within the literature-reported ranges for West Africa in 2017 (as per Figure 1 generated with data from [11,12] were retained. Specifically, model projected HIV prevalence of 0.5-6% in the general female population, 0.5-4% in the general male population, 15-48% in the brothel-based FSW group and 10-25% in the non-brothel based FSW group were kept as model fits and to reflect the HIV prevalence in West African in Figure 1. No fitting criteria were applied to the female 2+ group, male 2+ group or the brothel-based FSW clients and non-brothel based FSW client groups due to the provision of insufficient data for these subgroups.

### Correlation analysis

Among the projected model outcomes we assessed the correlation of different model parameters with the HIV prevalence among the general male and female population. We computed the partial rank correlation coefficient (PRCC) between the different model parameters and, for those where there was correlation, the statistical significance (p-value) of the correlation with male and female HIV prevalence. By doing this we characterised the linear relationship between the different parameter inputs and different HIV prevalence values that correspond to values within West Africa. This analysis was run in Stata v14. We considered the correlation in different HIV prevalence brackets: firstly assessing the full range of HIV prevalence in males (0.1-4%) and females (0.1-6%). Then, given the variation in the HIV prevalence amongst females and males in West Africa across the different 14 countries, we considered three HIV prevalence categories for females {0.1-2%, 2-3%, 3-6%} and males {0.1-2%, 2-3%, 3-4%}

## Results

Overall, for model projected HIV prevalence to be 0.1-4% in the males, 0.1-6% in females, 15-48% in the brothel-based FSW group and 10-25% in the non-brothel based FSW group, the model generated a total of 11,164 fits. Tables 2 and 3 contain the results of the correlation analysis between HIV prevalence among females and males in 14 West African countries and the model parameters. Figure 3(a)-(b) show the graphical representation of the results from Tables 2 and 3, with the inner ring in each Figure containing the results from Table 2 and the outer ring containing the results from Table 3.

**Table 2:**
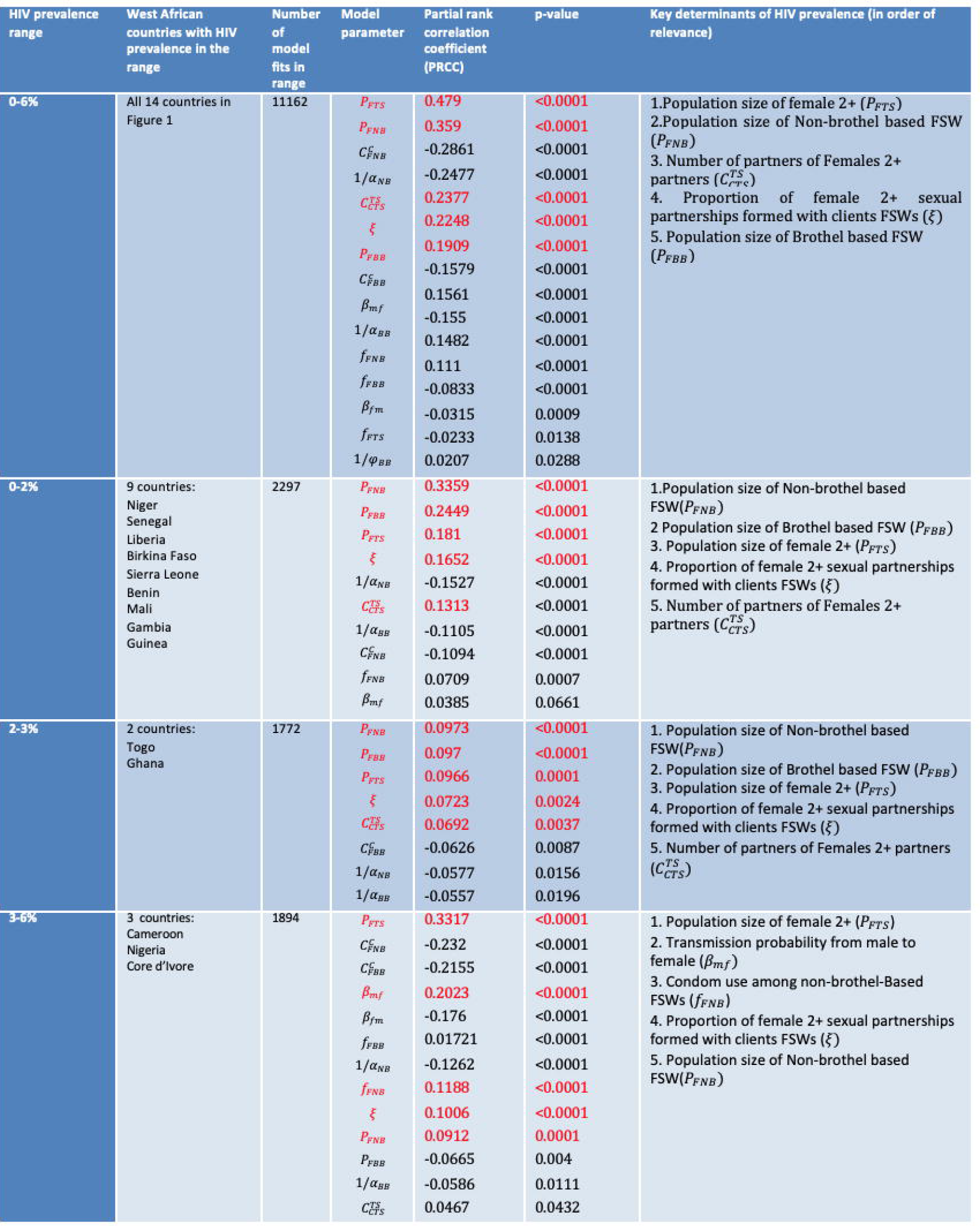
Results from the correlation analysis across different HIV prevalence ranges for the female population and across different West African countries that fit with these ranges. We project the impact and significance of the key model parameters.

**Table 3:** Results from the correlation analysis across different HIV prevalence ranges for the male population and across different West African countries that fit with these ranges. We project the impact and significance of the key model parameters.

| Prevalence range | West African countries with HIV prevalence in the range | Number of model fits in range | Input parameter | PRCC | p-value | Key determinants of HIV prevalence (in order of relevance) |
| --- | --- | --- | --- | --- | --- | --- |
| 0-4% | All 14 countries in Figure 1 | 11162 | $P_{FBB}$<br>$P_{FNB}$<br>$C_{CTS}^{TS}$<br>$1/\alpha_{NB}$<br>$P_{FTS}$<br>$\xi$<br>$1/\alpha_{BB}$<br>$C_{FBB}^C$<br>$\beta_{fm}$<br>$\beta_{mf}$<br>$C_{FNB}^C$<br>$f_{FBB}$<br>$f_{FNB}$<br>$f_{FTS}$ | 0.3646<br>0.3265<br>0.2431<br>-0.1964<br>0.1859<br>0.1762<br>-0.1701<br>0.1342<br>0.1015<br>-0.1011<br>-0.099<br>-0.089<br>0.0878<br>-0.07 | <0.0001<br><0.0001<br><0.0001<br><0.0001<br><0.0001<br><0.0001<br><0.0001<br><0.0001<br><0.0001<br><0.0001<br><0.0001<br><0.0001<br><0.0001<br><0.0001 | 1. Population size of Brothel-based FSWs ($P_{FBB}$)<br>2. Population size of Non-brothel based FSW ($P_{FNB}$)<br>3. Number of partners of Females 2+ partners ($C_{CTS}^{TS}$)<br>4. Population size of female 2+ ($P_{FTS}$)<br>5. Proportion of female 2+ sexual partnerships formed with clients FSWs ($\xi$)<br>6. Size of the group of clients to brothel-based FSWs ($C_{FBB}^C$)<br>7. Transmission probability from male to female ($\beta_{mf}$) |
| 0-2% | 12 countries:<br>Niger<br>Senegal<br>Ghana<br>Liberia<br>Birkina Faso<br>Sierra Leone<br>Benin<br>Mali<br>Gambia<br>Guinea<br>Togo<br>Core d'Ivoire | 2225 | $P_{FNB}$<br>$P_{FBB}$<br>$C_{CTS}^{TS}$<br>$1/\alpha_{NB}$<br>$\xi$<br>$\beta_{fm}$<br>$1/\alpha_{BB}$<br>$C_{FBB}^C$<br>$\beta_{mf}$<br>$P_{FTS}$<br>$1/\varphi_{BB}$ | 0.3063<br>0.2981<br>0.1578<br>-0.1248<br>0.1066<br>0.1041<br>-0.1036<br>0.0783<br>-0.0691<br>0.0648<br>-0.0503 | <0.0001<br><0.0001<br><0.0001<br><0.0001<br><0.0001<br><0.0001<br><0.0001<br>0.0002<br>0.0012<br>0.0023<br>0.0181 | 1. Population size of non-brothel-based FSWs ($P_{FNB}$)<br>2. Population size of Brothel based FSW ($P_{FBB}$)<br>3. Number of partners of Females 2+ partners ($C_{CTS}^{TS}$)<br>4. Proportion of female 2+ sexual partnerships formed with clients FSWs ($\xi$)<br>5. Transmission probability from male to female ($\beta_{mf}$) |
| 2-3% | 1 countries:<br>Cameroon | 2317 | $1/\alpha_{NB}$<br>$P_{FNB}$<br>$P_{FBB}$<br>$C_{CTS}^{TS}$<br>$f_{FNB}$ | -0.0797<br>0.062<br>0.0588<br>0.045<br>0.0466 | 0.0001<br>0.003<br>0.0048<br>0.0308<br>0.0323 | 1. Population size of non-brothel-based FSWs ($P_{FNB}$)<br>2. Population size of Brothel based FSW ($P_{FBB}$)<br>3. Number of partners of Females 2+ partners ($C_{CTS}^{TS}$)<br>4. Condom use among non-brothel based FSWs ($f_{FNB}$) |
| 3-4% | 1 country:<br>Nigeria | 2402 | $P_{FBB}$<br>$P_{FNB}$ | 0.0467<br>0.0391 | 0.0225<br>0.0504 | 1. Population size of Brothel-based FSWs ($P_{FBB}$)<br>2. Population size of Non-brothel based FSW ($P_{FNB}$) |

**Figure 3:**
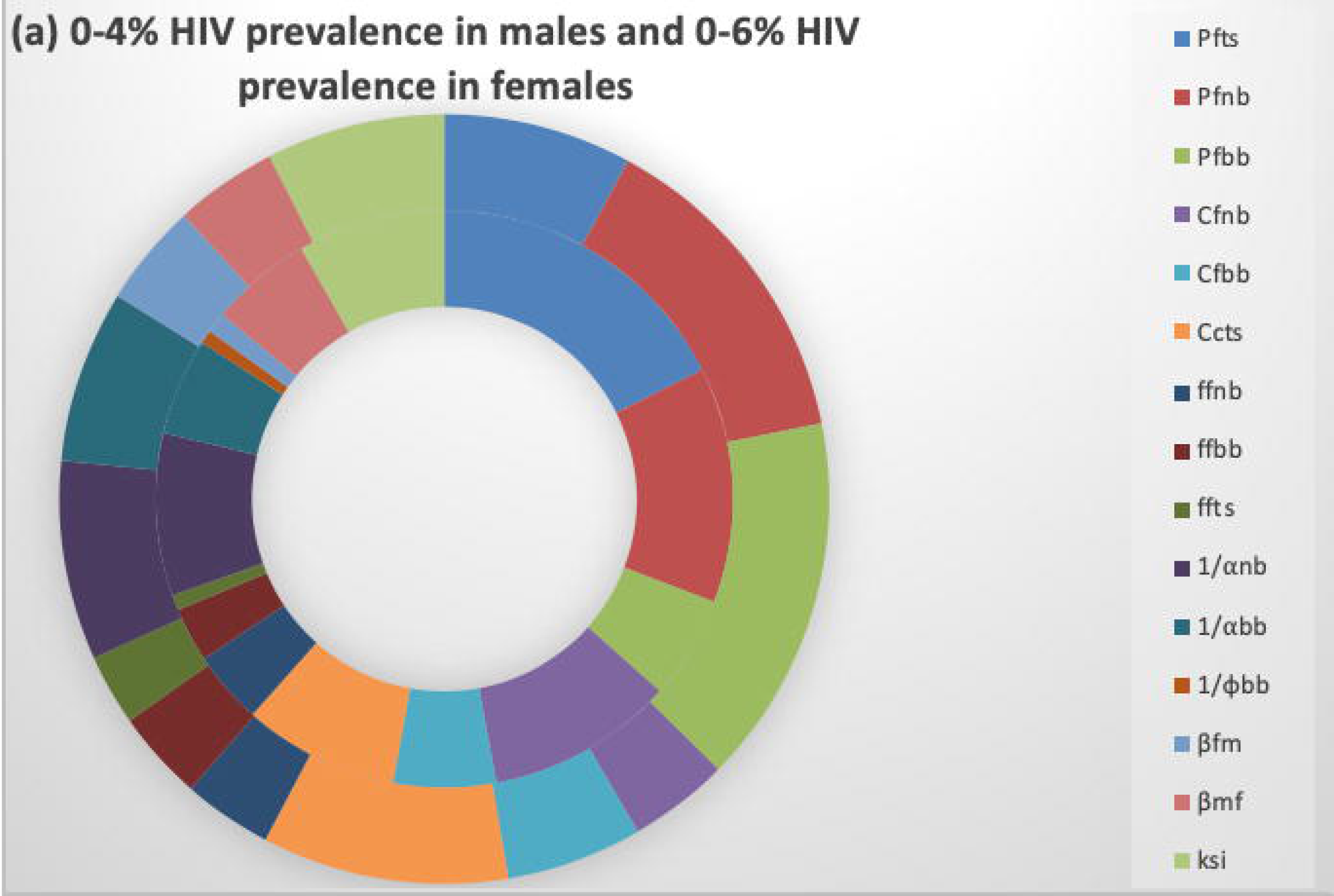
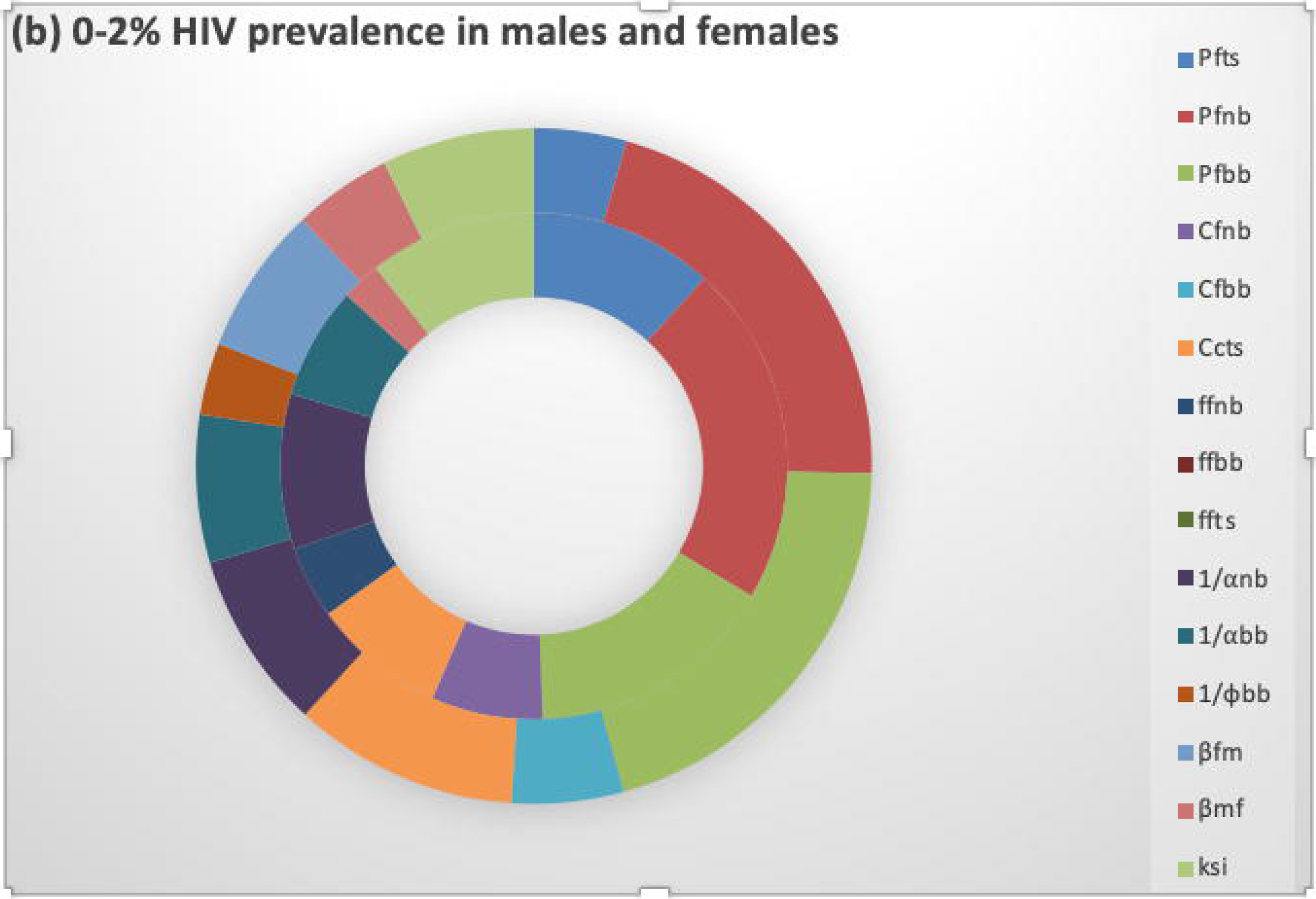
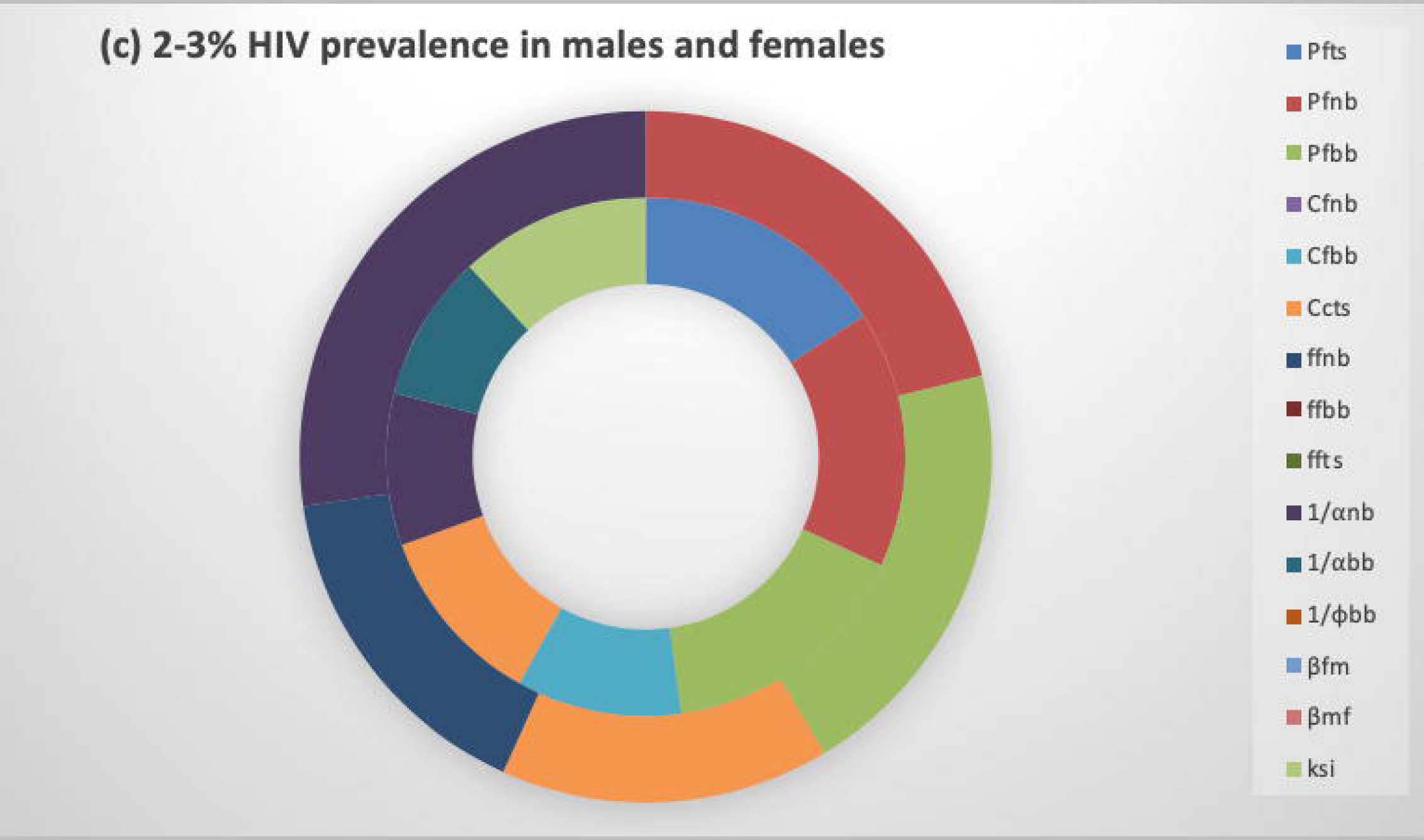
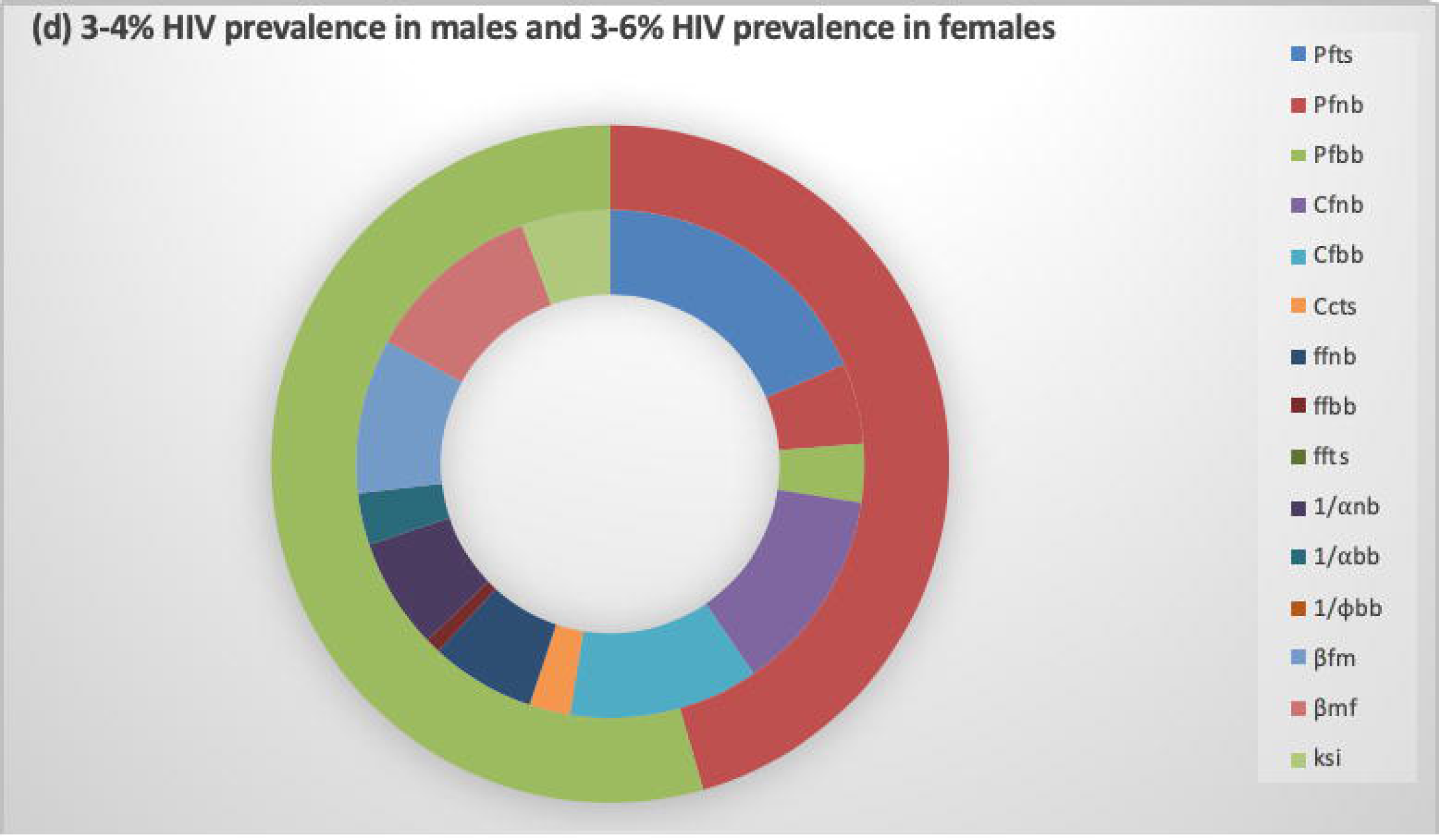
(a)-(d): Bar charts of the partial rank correlation coefficient (PRCC) between a selection of model parameters from Table 1 and the HIV prevalence in female and male general population across different HIV prevalence brackets. The absolute values of the PRCC are shown in Tables 2 and 3, and we only consider parameters that significantly influence the HIV prevalence (p-value <0.05).

### Key determinants of HIV prevalence in females

When we consider all countries collectively, where HIV prevalence among the female general population ranges from 0.1-6% (Table 2 and Figure 3(a) inner ring), the size of the population of the female 2+ group (*p*_*FTS*_) emerges as the most significant determinant of HIV prevalence among females. Of importance is also the number of sexual partners this group has 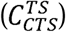, as well as the size of the non-brothel based sex-worker group (*p*_*FNB*_) and the parameter (*ξ*) that describes the proportion of sex acts females 2+ have with clients of FSWs (as opposed to their corresponding males 2+ group). The size of the brothel based FSW groups (*P*_*FBB*_) is also positively and significantly correlated with HIV prevalence among the females, highlighting the importance of the relative size of all sexually active female population groups.

In 9 out of the 14 countries, HIV prevalence among females is low (0.1-2%) and female HIV prevalence is most strongly correlated to the size of both brothel-based and non-brothel based FSW groups (*p*_*FNB*_ and *p*_*FBB*_) and to a lesser extent to the size of the group of females with 2+ partners (*p*_*FTS*_) (Figure 3(b)). However, the mixing pattern via the parameter (*ξ*) and the number of sexual partners of the female 2+ group 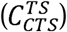 do remain positively correlated to female HIV prevalence here (Table 2; 2^nd^ row and Figure 3(b) inner ring). The results are similar in countries (2 out of 14) where HIV prevalence is between 2-3%, with the size of the FSW (*P*_*FNB*_, *P*_*FBB*_) groups most significantly correlated to HIV prevalence in females, and the size of the female 2+ group less important. Partnership numbers and duration of being a member of a higher-risk group (respectively (*ξ* and 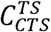) (Table 2; 3^rd^ row; Figure 3(c) inner ring), become more important factors in countries where HIV prevalence is slightly higher. For countries in which female general population HIV prevalence is within the highest prevalence range, 3-6% (3 out of 14 countries), the model indicates that the size of the female 2+ group (*P*_*FTS*_) is most strongly associated with HIV prevalence (Figure 3(d) inner ring). The parameter describing the proportion of partnerships females with 2+ partners form (*ξ*) with clients of FSWs, continues to be an important driver of HIV prevalence, while the size of the FSW groups becomes relatively less significant. This final observation is partly seen because the size of the FSW groups within the model are constrained, based on evidence from surveys.

### Key determinants of HIV prevalence in males

For males, HIV prevalence ranges from 0.1-4% across the14 countries. Our results show that the population size of the FSW groups (both brothel and non-brothel based; respectively *P*_*FBB*_ and *P*_*FNB*_) is the most important determinant of HIV prevalence for the male general population in West Africa (Figures 3(a)-(d) outer ring). The number of client partners that sex workers have 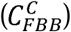 and the transmission probability of acquiring HIV as a female from a male partner (*β*_*mf*_) (Figure 3(a) outer ring) are important. The number of sexual partners of females 2+ 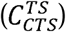 and the size of the population size of female 2+ (*P*_*FTS*_)are also positively corelated with male HIV prevalence but to a lesser extend (Table 3), as is the parameter (*ξ*) that describes the proportion of sex acts females 2+ have with clients of FSWs.

In West African countries with lower HIV prevalence among the male general population (0-2% in 12 out of 14 countries), we observe that the most relevant determinant of HIV prevalence is the population size of non-brothel-based FSWs (*P*_*FNB*_) and brothel based FSW (*P*_*FBB*_). In countries with higher HIV prevalence in the male general population (2-3% in 1 of 14 countries) the size of the FSWs groups (*P*_*FNB*_ and *P*_*FBB*_) remains important, with the number of partners of Females 2+ partners 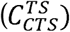 and the condom use among non-brothel based FSWs (*f*_*FNB*_) also correlated but to a lesser extent (Table 3 and outer ring of Figure 3(c)). Finally, in countries with higher HIV prevalence among the male general population (3-4%, 1 of 14 countries) only two parameters describing the size of the two FSW groups significantly affect the HIV prevalence in males (Table 3; last row and outer ring of Figure 3(d))).

## Discussion

Our results suggest that as female HIV prevalence increases across different West African countries, from 0.1-2% (in 9 out of 14 countries) through to 2-3% (2 out of 14 country) and then 3-6% (in 3 out of 14 country), the correlation between the parameters associated with the groups of female with 2+ partners and the HIV prevalence among females increases (results within the inner ring of Figure 3(a)-(d) and pictorially represented in Figure 4). Therefore, our results suggest that this group of females with 2+ partners, may have a gearing-type role in sustaining high HIV prevalence among females in West Africa.

**Figure 4:**
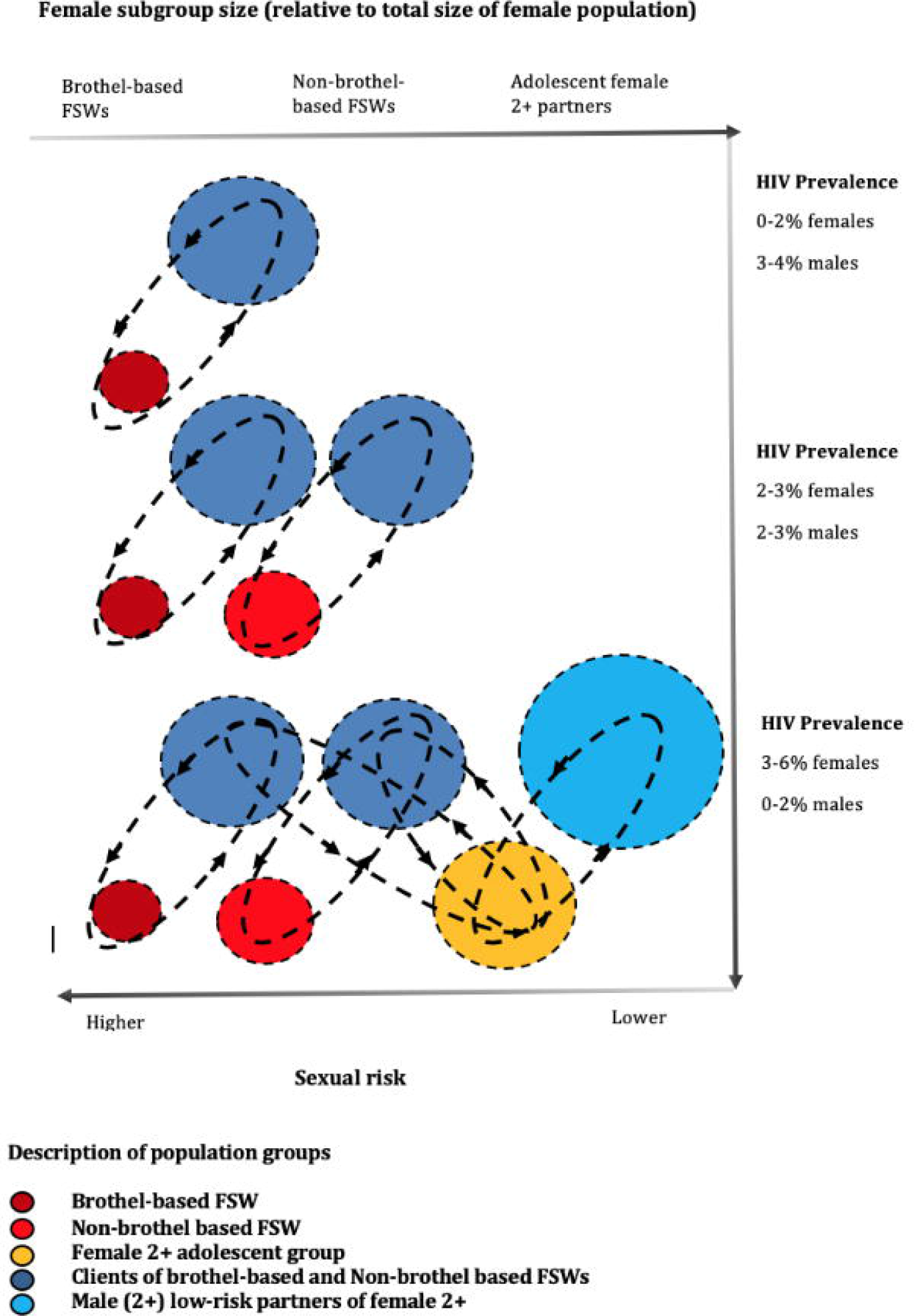
Conceptual idea of the “epidemic gearing” effect of HIV epidemics in West Africa (the black dashed lines represent the “gearing effect” of the epidemic, by representing the sexual network connections between risk groups in the population)

In contrast, as male HIV prevalence increases, from 0.1-2% (in 12/14 countries) through to 2-3% (1/14 country) and then 3-4% (in 1/14 country), the correlation between the parameters associated with the FSWs groups and the HIV prevalence among males increases (results within the outer ring of Figure 3(a)-(d) and pictorially represented in Figure 4). Therefore, our results show that the size of the FSW groups and their sexual activity may be more responsible for sustaining high HIV prevalence among males in West Africa.

Past studies have emphasised the importance of sex work and other vulnerable groups as key determinants of HIV prevalence [13] to sustain HIV prevalence of 0-3% among the general population [6] and demonstrated that the size of the FSW population (relative to the total population) is the most important determinant of HIV prevalence [14]. The findings from our study show that whilst (brothel and non-brothel) sex work is an important determinant of HIV prevalence in both females and males across the 14 West African countries, the size of the female 2+ group, also emerges as an important determinant of HIV prevalence, particularly for countries with higher prevalence levels and among the female HIV prevalence. For lower level epidemics in most countries, the size of FSW groups remains the key determinant in agreement with the findings of [14]. However, when stratifying the HIV epidemics into respective categories of HIV prevalence in both males and females, we provide evidence of a potential transitionary phrase within an epidemic which we depicted in Figure 4. This is an interesting hypothesis which potentially highlights a change in the nature of an HIV epidemic (perhaps more formally known as going from being a ‘concentrated’ to a ‘generalised’ epidemic, as characterised by UNAIDS), from one being more dependent on the size and sexual activity of female sex workers, to an epidemic in which the subgroup size and sexual activity of young females, with multiple partners, is of equal importance. This same transition is not seen when the analysis is repeated for males, which aligns more consistently with the theory of a larger population of male clients acting as a ‘bridge’ to female partners. Whilst traditionally in modelling analyses, these females were thought to be ‘long term’ steady partners, our analysis suggests that younger more vulnerable females could also act as a highly vulnerable group for acquiring new HIV infections, particularly if they are forming partnerships with men who are also clients of FSWs.

The findings from this analysis are important. Firstly, they confirm three previous findings: (a) that commercial sex work remains important for HIV transmission in West Africa, particularly in lower level epidemics and for sustaining HIV prevalence among the male general populations, in agreement with previous findings [14] (b) that when HIV prevalence is high amongst FSWs, a large proportion of HIV infections may be attributable to these groups alone and is in line with previous findings [15]; and (c) they support the hypothesis on the importance of behaviours that modify the risk of acquiring infection (e.g. fewer partner numbers, shorter durations in sex work and higher levels of condom use) as protective barriers against infection. Secondly, our results are in support of the idea of a type of “*epidemic gearing*” effect depicted in Figure 4. This will imply that, for the epidemic to have the propensity to grow, it needs to be effectively “geared up”, firstly, by smaller high-sexual activity groups of FSWs, and then secondly, through the larger subgroup of adolescent females 2+, which act as an additional larger “cog” in the epidemic chain enabling the epidemic to achieve higher-levels of HIV prevalence (Figure 4). Here the concept of behavioural heterogeneity and the importance of approaches that seek to identify high-risk individuals and understanding the structure of sexual networks emerge as important.

Although our results are important, for epidemics in countries where HIV prevalence is high (e.g. higher than 4% in West Africa), transmission pathways may require a more advanced understanding of the underlying factors driving the epidemic. We strongly support the collection of more behavioural data to better understand and inform these pathways, and advocate for closer collaborations between modellers and social scientists, so that mathematical model structures better reflect the true dynamics of HIV epidemics.

Our analysis does have several limitations. Estimates on the percentage of adolescent females reporting 2+ partners is mostly extracted from the DHS data. Whilst there is some comparability internally and between countries, typically questions on sexual behaviours are prone to under-reporting, especially amongst women. All results should be interpreted with caution, and the nature of the findings are partly a consequence of the model’s structure, although we believe this to be a more useful representation of sexual networks and transmission pathways, supported by evidence both from the modelling and social science literature.

Population data from DHS surveys suggests that the percentage of adolescent females with multiple partners, may comprise up to 9% of the total female population, with other studies reporting higher percentages [16-18]. This modelling study, shows the importance of the size and sexual activity of the female 2+ group and more broadly the need to assess and understand behaviours that shape sexual network structures as well as those that modify the risks of acquiring infection, in determining variations in HIV prevalence. However, to date, very few mathematical models of HIV transmission in the general population explicitly include or recognise the importance of an adolescent female 2+ group, despite the high levels of incidence in this population. In addition, more accurate data on the estimated subgroup size for high-risk groups of female sex workers is often absent from modelling studies, despite demonstrating here the fundamental importance of this.

In our correlation analysis we have chosen to use PRCC rather than other correlation coefficients, as we believe that it provides good insight on the global sensitivity of the system and projects the parameters that are most influential (or significant) even if other parameters are simultaneously perturbed. Other correlation analysis (e.g. Pearson correlation) are more based on local sensitivity calculations and can provide insight into small perturbations around the key parameters, but can often ignore the impact of large perturbations, something we wanted to include. Future analysis will consider contrasting different correlation measures and discuss the differences in the findings.

Our findings have the potential to have important implications for future policy. The UNAIDS definition for a concentrated epidemic is one in which HIV prevalence is less than 1% in the general population and over 5% prevalence in key risk groups such as FSWs, with the caveat being that no subpopulation is fully self-contained and these thresholds should be interpreted with caution [19]. However, here we demonstrate the dangers of such rigid definitions, by highlighting the subtle maturity that may occur in epidemics from those driven predominantly by commercial sex work, to those in which both the role of commercial sex and the size and sexual behaviours of other populations is important. The social science literature should be a greater source of information, highlighting in particular the vulnerability of young females [20,21].

Finally, despite much focus on interventions being towards reducing the biological probability of transmission, our findings suggest that programmes which result in fewer women practicing sex work and fewer young females engaging in higher-risk partnerships, could play a key role in reducing the size of the HIV epidemics in West Africa. In the future, sex work may become less prevalence as women become more empowered with better education, jobs and earning potential as African countries continues to develop. Future policies on HIV should focus on treatment, prevention, but also key aspects of women’s development, to ensure they are able to make the best most informed decisions.

## Data Availability

The data and the mathematical model are available from the corresponding author on reasonable request.

## Acknowledgements

We would like to thank Prof Peter Vickerman (University of Bristol) and Dr Kate Mitchell (Imperial College London) for their early contributions to this work, to Prof Timothy Hallett (Imperial College London) and Dr Deirdre Hollingsworth (Warwick University) for their contributions the methodology of the correlation analysis and to Prof Graham Medley (LSHTM) for his insightful contribution to discussion of the interpretation of the findings from this study.

## Supplementary Material

Appendix A: Description of the novel mathematical model for HIV transmission in West Africa

Supplementary Table S1: Model parameters and their values

